# Contribution of Mendelian disorders in a population-based pediatric neurodegeneration cohort

**DOI:** 10.1101/2022.03.15.22271876

**Authors:** Rose Guo, Frank D. Mentch, Dong Li, Erin E. Will, Rebecca C. Ahrens-Nicklas, Elizabeth J. Bhoj

**Affiliations:** Division of Human Genetics, Children’s Hospital of Philadelphia, Philadelphia, Pennsylvania, USA; Center for Applied Genomics, Children’s Hospital of Philadelphia, Philadelphia, Pennsylvania, USA; Division of Genomic Diagnostics and Department of Pathology, Children’s Hospital of Philadelphia, Philadelphia, Pennsylvania, USA

**Keywords:** Regression, Pediatric Neurology, Medical Genetics, Rett syndrome, mitochondrial disease, Neuroscience, Biobanking

## Abstract

**Objectives:** To evaluate Mendelian causes of neurodegenerative disorders in a cohort of pediatric patients as pediatric neurodegenerative disorders are a rare, diverse group of diseases. As molecular testing has advanced, many children can be diagnosed, but the relative contribution of various disorders is unclear.

**Study Design:** Patients enrolled in the Center for Applied Genomics (CAG) Biobank at the Children’s Hospital of Philadelphia with neurodegenerative symptoms were identified using an algorithm that consisted of including and excluding selected ICD9 and ICD10 codes. A manual chart review was then performed to abstract detailed clinical information.

**Results:** Out of approximately 100,000 patients enrolled in the CAG Biobank, 76 had a neurodegenerative phenotype. Following chart review, 7 patients were excluded. Of the remaining 69 patients, 42 had a genetic diagnosis (60.9%) and 27 were undiagnosed (39.1%). There were 32 unique disorders. Common diagnoses included Rett syndrome, mitochondrial disorders and neuronal ceroid lipofuscinoses.

**Conclusions:** The disorders encountered in our cohort demonstrate the diverse diseases and pathophysiology that contribute to pediatric neurodegeneration. Establishing a diagnosis often informed clinical management, although curative treatment options are lacking. Many patients who underwent genetic evaluation remained undiagnosed, highlighting the importance of continued research efforts in this field.

## Introduction

Pediatric neurodegenerative disorders are a diverse group of diseases that cause varying neurologic symptoms, such as developmental delay, regression, intellectual disability, seizures, ataxia and weakness. Abnormalities are often present on brain MRI. Although rare with an incidence of 0.6 in 1,000 live births^1,2^, the progressive nature and lack of curative treatments for many of these disorders leads to significant morbidity and a high mortality rate^3^. Some pediatric neurodegenerative disorders have known genetic etiologies and are increasingly being better characterized. Many novel monogenic causes have been discovered in recent years as genetic testing methods have advanced, but the relative contribution of various disorders is unclear. Knowledge of the molecular mechanisms has informed clinical management, however, treatment options are limited. Unfortunately, pediatric patients with neurologic symptoms who undergo clinical genetic evaluations only receive a molecular diagnosis 35% of the time^4^.

Studying the mechanisms of pediatric neurogenerative disorders can additionally help elucidate the pathology of adult-onset neurodegenerative disorders. There appear to be common pathways involving lysosomal dysfunction, mitochondrial dysfunction^5,6^ and autophagy defects^7^ that span all age groups. For example, neuronal ceroid lipofuscinoses (NCLs) are a group of neurodegenerative lysosomal storage disorders caused by variants in 14 *CLN* genes, respectively named *CLN1-14*^8^. Loss of function variants in *CLN12* have been shown to cause an autosomal recessive juvenile-onset Parkinson dementia called Kufor-Rakeb syndrome, and have also been implicated in the pathophysiology of adult-onset frontotemporal dementia^9^. Another example is the link between Gaucher disease and Parkinson disease. Gaucher disease is an autosomal recessive lysosomal storage disorder caused by variants in the *GBA* gene. Parkinsonian symptoms have been seen in a large proportion of Gaucher disease patients in the 4^th^ to 6^th^ decade of life^10^ and conversely, many individuals with Parkinson disease are found to be heterozygous for pathogenic *GBA* variants^11^, suggesting a common neurodegenerative pathway.

Here, we describe Mendelian causes of neurodegenerative disorders encountered in a cohort of pediatric patients to demonstrate the breadth of potential diagnoses and to examine how a genetic diagnosis can impact clinical management.

## Methods

A cohort of pediatric patients with a neurodegenerative phenotype was generated using an algorithm to pull patients enrolled in the Center for Applied Genomics (CAG) Biobank at the Children’s Hospital of Philadelphia. The cohort was selected in a multi-step process and included both automated and manual steps. First, we used 44 ICD9 and ICD10 codes specific for neurodegeneration and those likely related to neurodegeneration (Table 1) were used to create an initial cohort. Then, ICD codes for infectious disease and autoimmune disease (Supplemental Tables 1 and 2) were used to filter out patients from this initial group who were not of interest.

**Table 1:** Initial Neurodegeneration Inclusion ICD9 and ICD10 Codes Families, Encompassing 44 Total ICD9 and ICD10 codes.

| Condition | ICD type | Code(s) |
| --- | --- | --- |
| cerebral_degeneration | ICD9 | 330% \| 331% |
| cerebral_degeneration | ICD10 | G30% \| G31% \| G32% |

The next step was a review of all the specific diagnosis descriptions to further filter the cohort. Patients were excluded based on diagnosis descriptions rather than codes because some of the codes included both desired and undesired conditions.

The resulting list of patients was then submitted for manual chart review. Electronic health records, linked to the Biobank, were reviewed including all available diagnostic testing, imaging studies, and clinical notes. Detailed clinical information was abstracted for patients with a diagnosis.

This study was conducted with approval from the Institutional Review Board of the Children’s Hospital of Philadelphia.

## Results

Out of approximately 100,000 pediatric patients enrolled in the CAG Biobank, 76 were identified to have a neurodegenerative phenotype (Figure 1). Following manual chart review, seven patients were excluded for having secondary neurologic symptoms due to other medical issues (one with a cavernous brainstem malformation, one with cerebral infarcts due to sickle cell disease, one with medulloblastoma, one with meningitis, one with brain injury due to nonaccidental trauma) and two for complete lack of neurologic symptoms. Of the remaining 69 patients, 42 had a genetic diagnosis (60.9%) and 27 were undiagnosed (39.1%). Of those diagnosed, there were 32 unique disorders (Table 2 and Supplemental Table 3). Common diagnoses were monogenic and included Rett syndrome, mitochondrial disorders (such as Leigh syndrome), neuronal ceroid lipofuscinoses, X-linked adrenoleukodystrophy, and Aicardi Goutieres syndrome. One patient (patient 9) had variants in two different genes identified, leading to a dual diagnosis of *AFG3L2*-related mitochondrial disease and spastic paraplegia type 7. Testing methodologies included karyotype, chromosomal SNP microarray, targeted single gene analysis, gene panels, mitochondrial genome analysis and whole exome sequencing. 38/40 (95%) were diagnosed through clinical testing, while 2/40 (5%) were diagnosed on a research basis. Testing methodology was unknown for two cases. Establishing a genetic diagnosis led to a change in clinical management through surveillance of known associated complications, medication management, and/or inclusion in a clinical trial in 39/42 (92.8%) cases. A total of 8/69 (11.5%) patients were deceased, 6/42 (14.2%) with a diagnosis and 2/27 (7.4%) undiagnosed. For all of the diagnosed cases it was felt that their molecular diagnosis completely explained their clinical findings.

**Figure 1:**
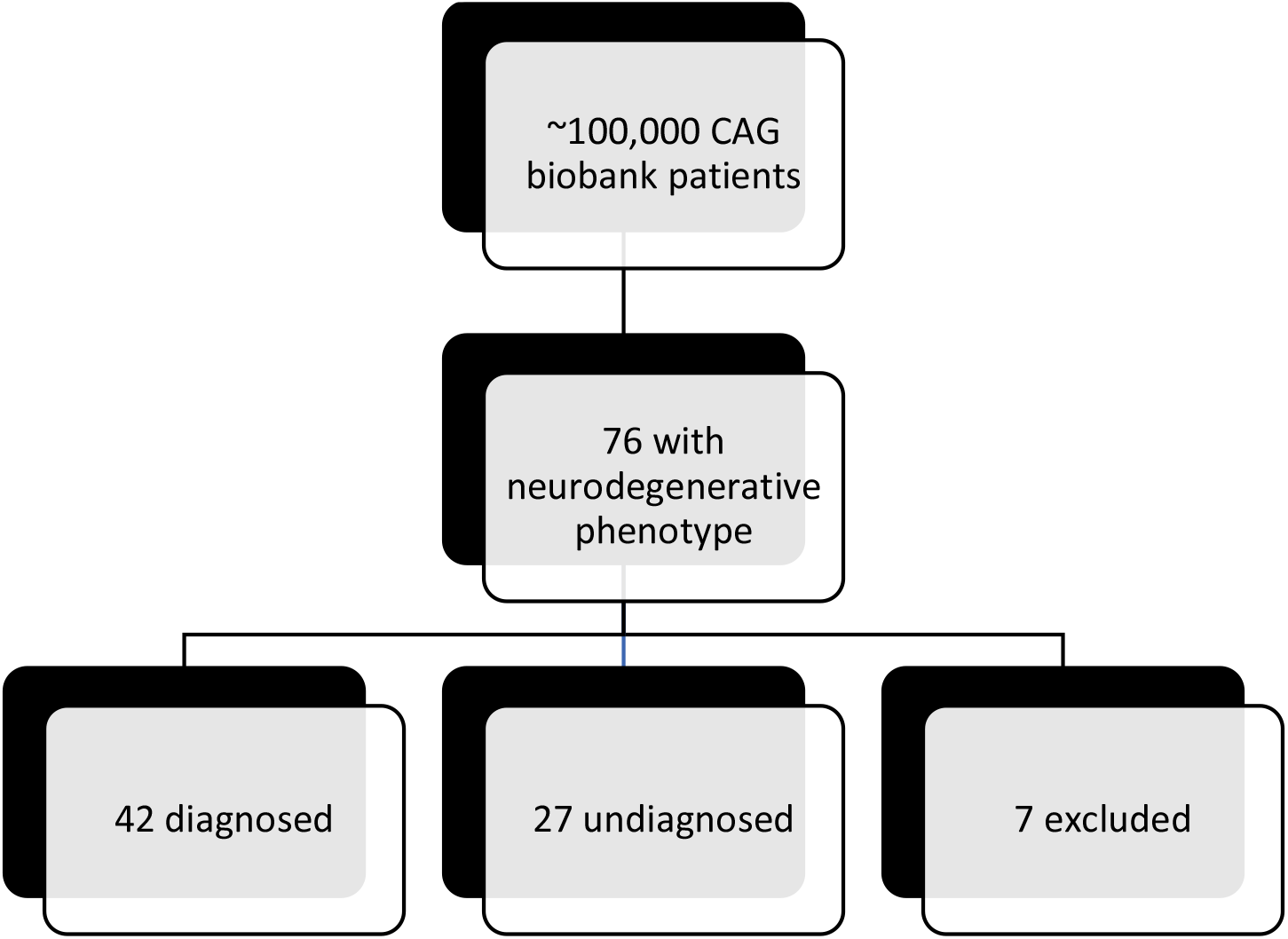
Stratification of Pediatric Neurodegeneration Cohort.

**Table 2.** Mendelian disorders encountered in the diagnosed cohort.

| Disorder | Number of Patients |
| --- | --- |
| Rett syndrome | 6 |
| Leigh syndrome (total) | 6 |
| Pyruvate dehydrogenase deficiency | 1 |
| • Complex I deficiency | 3 |
| • Complex V deficiency | 2 |
| Neuronal Ceroid Lipofuscinoses (total) | 4 |
| • Type 1 (NCL1), infantile | 1 |
| • Type 1 (NCL1) | 1 |
| • Type 2 (NCL2) | 1 |
| • Juvenile (JNCL) | 1 |
| X-linked Adrenoleukodystrophy | 2 |
| Aicardi Goutieres syndrome | 2 |
| ADCY5-related disorder | 1 |
| GNB1-related disorder | 1 |
| KIF1A-related disorder | 1 |
| SPTAN1-related disorder | 1 |
| TUBA1A-related disorder | 1 |
| SLC39A8-Congenital Disorder of Glycosylation (CDG) | 1 |
| Single large scale mtDNA deletion syndrome | 1 |
| FBXL4-related mitochondrial disease | 1 |
| Combined AFG3L2-related mitochondrial disease + Spastic paraplegia type 7 | 1 |
| Mitochondrial disorder not otherwise specified | 1 |
| Infantile Neuroaxonal Dystrophy | 1 |
| Galloway-Mowat syndrome | 1 |
| Noonan syndrome | 1 |
| Alexander disease | 1 |
| Sandhoff disease (GM2 Gangliosidosis) | 1 |
| Pelizaeus-Merzbacher disease | 1 |
| Metachromatic Leukodystrophy | 1 |
| Autosomal recessive spastic ataxia of Charlevoix-Saguenay (ARSACS) | 1 |
| Cono-renal syndrome | 1 |
| Gaucher Type 1 | 1 |
| Fumarase deficiency | 1 |
| 1p36 deletion | 1 |
| Total | 42 |

## Discussion

The disorders encountered in our cohort demonstrate the diverse, yet common pathophysiologic pathways in pediatric neurodegenerative disease that involve primary neuronal function as well as lysosomal, mitochondrial and/or autophagy defects that secondarily impact neuronal health. Some are well-known, while others are novel single gene disorders and highlight differential diagnoses and testing methods that should be considered for those undergoing genetic evaluation for a neurodegenerative disorder. While the majority were monogenic diagnoses, one case (patient 9) had a dual diagnosis of *AFG3L2*-related mitochondrial disease and spastic paraplegia type 7 which was identified through clinical whole exome sequencing. The three most commonly seen disorders in our cohort were Rett syndrome, Leigh syndrome, and neuronal ceroid lipofuscinoses. Establishing a genetic diagnosis allows for prognostication, surveillance recommendations based on known associated health complications and even potential medications or therapeutic interventions if available as demonstrated in the following examples of our most common diagnoses.

The most frequent diagnosis, Rett syndrome, is characterized by normal development in the first 6-18 months of life followed by a period of stagnation, then rapid onset of neurodevelopmental regression^12^. Additional features such as seizures and ataxia may be seen. It is an X-linked disorder arising from variants in *MECP2* which disrupt methylation and/or regulation of transcription. Although there is no curative treatment, there are a variety of known associated medical complications, including QTc prolongation and osteopenia, that require ongoing surveillance and medical management.

The second most common diagnosis, mitochondrial disorders, are a large, expanding group of diseases, many of which have neurologic features as the initial presentation in childhood. While complex and often difficult to diagnose, some commonalities in clinical features have allowed for categorization, such as Leigh syndrome. Leigh syndrome is characterized by subacute relapsing and necrotizing encephalopathy causing neurologic stagnation or regression followed by eventual progressive decline combined with classic imaging findings of bilateral symmetric basal ganglia and/or brainstem lesions^13, 14^. There are over 95 combined nuclear and mitochondrial genomic etiologies of Leigh syndrome to date.

Management guidelines involve surveillance for neurologic, ophthalmologic, audiologic and cardiologic complications and supportive treatment with mitochondrial cocktail to support the mitochondrial respiratory chain. Avoidance of certain anesthetics and other medications can prevent secondary complications. During periods of acute illness, close monitoring is warranted to assess for decompensation. If an acute metabolic stroke is suspected, intravenous arginine therapy can be considered in appropriate circumstances as seen in our patient cohort^15^. For several mitochondrial disorders, clinical trials are under development. For example, a trial of dichloroacetate (NCT02616484) is ongoing to investigate its role as a potential treatment in genetic mitochondrial disease, particularly in pyruvate dehydrogenase complex deficiency^16^.

And the third most common diagnosis, neuronal ceroid lipofuscinoses (NCLs), are a group of lysosomal storage disorders that commonly cause neurodevelopmental regression and dementia in children^17^. There are currently 14 unique molecular NCL subtypes (CLN1-CLN14). Onset is variable, ranging from severe infantile presentations to milder adult disease. Many are progressive in nature and can cause rapid decline with high mortality. Not only is diagnosis important for prognostication, but there is also a growing number of treatment and clinical trial options available for the NCL disorders. For example, cerliponase alfa, a brain-directed enzyme replacement therapy, was recently approved by both the FDA and the EMA for the treatment of CLN2 disease. While none of these treatment options allows for complete reversal of the disease, early diagnosis is crucial if a disease-modifying treatment is available to halt or slow progression.

Limitations of our study include a potentially skewed population of children that are enrolled in the CAG biobank. Patients who were receiving a blood draw as part of routine care were approached for enrollment into the CAG, allowing for the creation of a theoretically unbiased biobank. However, enrollment only occurred on the CHOP main campus and no outpatient sites were included where a larger proportion of children without underlying medical issues potentially may have blood draws performed. Additionally, the algorithm used to identify the pediatric neurodegenerative phenotype relied on using specific ICD codes, so if these were not accurately entered into the patient’s medical chart, their phenotype may not be captured.

The overall mortality rate in our cohort was 11.5% (14.2% in the diagnosed group, 7.4% in the undiagnosed group), emphasizing the high mortality associated with pediatric neurodegenerative disorders, likely due to their progressive nature and lack of curative treatment options. A large number of cases also remained undiagnosed (39.1%), although this number was lower than nondiagnostic rates of 65% previously reported in the literature^4^. Our population likely had a high diagnostic rate due to careful phenotyping by trained geneticists, availability of both clinical and research-based testing and reanalysis capabilities. Notably, 24% (6/25) of the undiagnosed patients had exome sequencing, while 33% 14/42 of the diagnosed patients had exome sequencing. It is very possible that more of the undiagnosed patients would have benefitted from clinical exome sequencing.

Much remains to be discovered in the field of pediatric neurodegeneration not only to improve mortality rate, but also to better understand the pathophysiology behind these disorders to increase the diagnostic yield and provide targeted treatment options. Ongoing research efforts are of critical importance. In our cohort, 5% of patients received diagnoses through research testing and if not for these opportunities, would otherwise remain undiagnosed. Given the many undiagnosed patients in the cohort we suggest that novel gene-disease discovery should include an integrated genomic, transcriptomic and metabolomic analysis given the vast mechanistic possibilities.

## Data Availability

All data produced in the present work are contained in the manuscript

## Data Availability

All data are available within the paper.

## Acknowledgements

See Disclosures.

## Author Contributions

conceptualization: RG, RAN, EB; Data curation: DL, RG, RAN, EB, EW; Formal analysis: DL, RG, RAN, EB, FM; Investigation: RG, RAN, EB, FM; Writing– original draft: RG; Writing–review & editing: DL, RG, RAN, EB, FM.

## Ethics Declaration

This study was conducted with approval from the Institutional Review Board of the Children’s Hospital of Philadelphia. Patients and families provided written consent for enrollment in the Center for Applied Genomics (CAG) Biobank at the Children’s Hospital of Philadelphia. Patient information was de-identified for this publication.

## Figures

**Supplemental Table 1:** Infectious Disease Exclusion Codes.

| Disease | ICD9 Code(s) |
| --- | --- |
| H Influenza | 038.41% \| 041.5% \| 320.0% \| 482.2% |
| meningococcal | 036.0% \| 036.1% \| 036.2% \| 036.3% \| 036.4% \| 036.81% \| 036.82% \| 036.89% \| 036.9% |
| pneumococcal | 038.2% \| 320.1% \| 481% \| 567.1% |
| staphylococcal | 038.10% \| 038.11% \| 038.12% \| 038.19% \| 041.10% \| 041.11% \| 041.12% \| 041.19% \| 482.40% \| 482.41% \| 482.42% \| 482.49% \| v02.53% \| v02.54% \| v12.04% |
| e coli | 008.0% \| 038.42% \| 041.4% \| 482.82% \| V01.83% |
| rhinovirus | %rhinovir% \| 079.3% |
| Rsv | 079.6% \| 466.11% \| 480.1% |
| adenovirus | 008.62% \| 049.1% \| 079.0% \| 480.0% |
| enterovirus | 008.67% \| 047% |

**Supplemental Table 2:** Autoimmune Disease Exclusion Codes.

| Disease | ICD type | ICD Code(s) |
| --- | --- | --- |
| ARTER | ICD9 | 437.4% \| 446% \| 446.0% \| 446.5% \| 446.7% \| 447.6% |
| ARTER | ICD10 | I67.7% \| I68.2% \| I77.6% \| I79.1% \| M30% \| M30.0% \| M30.2% \| M30.8% \| M31.4% \| M31.5% \| M31.6% \| M31.7% |
| ARTH | ICD9 | 714% \| 714.0% \| 714.1% \| 714.2% \| 714.3% \| 714.30% \| 714.31% \| 714.32% \| 714.33% \| 714.4% \| 714.8% \| 714.81% \| 714.89% \| 714.9% |
| ARTH | ICD10 | M05% \| M06% \| M08.0% \| M08.2% \| M08.3% \| M08.4% \| M08.8% \| M08.9% |
| AST | ICD9 | 493% |
| AST | ICD10 | J45% |
| BEHC | ICD9 | 136.1% \| 711.2% \| 711.20% \| 711.21% \| 711.22% \| 711.23% \| 711.24% \| 711.25% \| 711.26% \| 711.27% \| 711.28% \| 711.29% |
| BEHC | ICD10 | M35.2% |
| CD | ICD9 | 555% |
| CD | ICD10 | K50% |
| CNJ | ICD9 | 372.0% \| 372.1% |
| CNJ | ICD10 | H10.1% \| H10.4% |
| CTD | ICD9 | 710.8% |
| CTD | ICD10 | M35.1% |
| CVID | ICD9 | 279.06% |
| CVID | ICD10 | D83% |
| DERM | ICD9 | 691% |
| DERM | ICD10 | L20% |
| EOS | ICD9 | 530.13% |
| EOS | ICD10 | K20.0% |
| FOOD | ICD9 | V15.01% \| V15.04% \| V15.08% |
| FOOD | ICD10 | Z91.010% \| Z91.013% \| Z91.041% |
| GDPAST | ICD9 | 446.21% |
| GDPAST | ICD10 | M31.0% |
| GLMN | ICD9 | %glomerulonephritis% |
| HEP | ICD9 | 571.42% |
| HEP | ICD10 | K75.4% |
| HSP | ICD9 | 287.0% |
| HSP | ICD10 | %D69.0% |
| JIA | ICD9 | 714.2% %syst% \| 714.3% %syst% \| 714.30AZ \| 714.30BA |
| JIA | ICD10 | M08.20% |
| MCD | ICD9 | 581.3% |
| MCD | ICD10 | N04% |
| MYOS | ICD9 | 710.3% \| 710.4% |
| MYOS | ICD10 | M33% \| M33.0% \| M33.00% \| M33.01% \| M33.02% \| M33.09% \|<br>M33.1% \| M33.10% \| M33.11% \| M33.12% \| M33.19% \|<br>M33.2% \| M33.20% \| M33.21% \| M33.22% \| M33.29% \|<br>M33.9% \| M33.90% \| M33.91% \| M33.92% \| M33.99% \|<br>M36.0% |
| OTM | ICD9 | 381.1% |
| OTM | ICD10 | H65.2% |
| PAN | ICD9 | 446% |
| PAN | ICD10 | M30% |
| PBC | ICD9 | 571.6% |
| PBC | ICD10 | K74.3% \| K74.5% |
| PSOR | ICD9 | 696.0% \| 696.1% |
| PSOR | ICD10 | L40% |
| PULMF | ICD9 | 515% |
| PULMF | ICD10 | J84.10% \| J84.9% |
| PYODERM | ICD9 | 686.0% \| 686.00% \| 686.01% \| 686.09% |
| PYODERM | ICD10 | L08.0% \| L08.81% \| L88% |
| RHN | ICD9 | 477% |
| RHN | ICD10 | H65.2% |
| SARC | ICD9 | 135% |
| SARC | ICD10 | D86% |
| SIN | ICD9 | 473% |
| SIN | ICD10 | J32% |
| SLE | ICD9 | 710.0% |
| SLE | ICD10 | M32.1% \| M32.8% \| M32.9% |
| SSS | ICD9 | %SJOEGREN% \| %Sjogren% \| 517.2% \| 710.1% \| 710.1% \|<br>710.2A% \| 710.2AR% \| 710.2B% \| 710.2K% |
| SSS | ICD10 | M34% \| M34.0% \| M34.1% \| M34.8% \| M34.81% \| M34.89% \|<br>M34.9% \| M35.0% \| M35.8% \| M35.9% |
| Tymp | Surgery | 00126.99% \| 69405% \| 69433% \| 69436% |
| UC | ICD9 | 556% |
| UC | ICD10 | K51% |

**Supplemental Table 3: Detailed Description of Diagnosed Patients**

See Excel sheet

